# Rapid functional remodeling of the targeted contralesional hemisphere induced by one week of noninvasive closed-loop neurofeedback guides motor recovery in post-stroke patients with chronic motor impairment: a phase I trial

**DOI:** 10.1101/2024.06.11.24308643

**Authors:** Kenichi Takasaki, Seitaro Iwama, Fumio Liu, Miho Ogura-Hiramoto, Kohei Okuyama, Michiyuki Kawakami, Katsuhiro Mizuno, Shoko Kasuga, Tomoyuki Noda, Jun Morimoto, Meigen Liu, Junichi Ushiba

## Abstract

Post-stroke hemiplegia of the upper extremities continues to pose a significant therapeutic hurdle. Contralesional uncrossed corticospinal pathways (CST) are involved in the recovery processes. Here, we test the feasibility, safety, and preliminary efficacy of targeted upregulation of uncrossed CST excitability using self-modulation training of cortical activities. In this phase I trial, eight individuals with persistent (1-8 years) severe post-stroke motor disability, who did not respond to conventional rehabilitation, voluntarily actuated their affected shoulder using a closed-loop, noninvasive digital connection, bridging the contralesional motor cortex (M1) and an exoskeleton robot. While patients attempted to elevate the affected arm, scalp electroencephalogram signals over the contralesional M1 were processed online to provide them with feedback on induced excitability change. Extensive neurophysiological examinations were conducted to evaluate the intervention effect using whole-head EEG and magnetic stimulations. The digital connection reconstructed neural pathways, allowing arm elevation contingent on voluntary upregulation of contralesional M1 excitability without any adverse effects. As evidenced by clinical score increases and neurophysiological examinations, seven days of consecutive system use resulted in rapid, sustained, and clinically significant improvement in motor function when removed from the system and promoted contralesional M1 functional remodeling. This closed-loop system is safe, feasible, and a promising intervention that recruits intact neural resources to allow patients to recover upper-extremity motor abilities. Larger clinical trials are needed to confirm these results.

## Main

Impairment of upper-extremity motor abilities after a stroke is highly treatment-resistant, and its disease burden on patients and the social economy remains a global clinical challenge. According to meta-analysis^1^, hemiplegic upper extremities in patients with mild to moderate motor disability can be improved by constraint-induced movement therapy, bilateral upper limb training, electromyographic (EMG) biofeedback, and robot-assisted training. These treatments aim to improve the contra- or ipsilesional corticospinal pathways but are not specific to either pathway due to the nature of behavioral training.

For patients with severe disability, whose residual motor functions do not allow extensive behavioral training, repetitive mental rehearsal of affected extremities movements has been used as a motor imagery therapy. However, the open-loop therapy in which patients explore mental strategies without feedback contributes to inconsistent therapeutic effects due to the heterogeneity of instruction and lack of monitoring methods for performance^2^. To overcome this limitation, recently proposed closed-loop approaches using functional neuroimaging techniques, including scalp electroencephalogram (EEG), aided this therapy by monitoring the induced neural excitability to provide patients with sensory feedback in a brain state-dependent manner^3,4^. Moreover, because the closed-loop approach enables flexible allocation of neural pathways recruited during the therapy, it could be optimized by targeting intact neural pathways that are directly connected to end-effector muscles in the affected side^5^.

To maximize the clinical efficacy of the closed-loop therapy, there is a need for the target selection of neural structures recruited during training. If flexible selection of therapeutic targets is achieved in a noninvasive manner, the focal closed-loop training would link attempted movements with the recruitment of a specific neural activity pattern in a context-specific manner^6^. Specifically, recruiting additional neural resources connected to the affected muscles, that is promoting functional neural remodeling of contralesional activity, can lead to motor recovery in arm movement involving proximal muscles bilaterally innervated by corticomotor pathways^7^. However, there are no methods to noninvasively select which pathways to recruit during the targeted movement.

In this study, we report our experience with treating eight chronic, severe post-stroke hemiplegic patients over a 7-day period of focal brain-machine interaction training using a noninvasive closed-loop connection between contralesional sensorimotor activities and an exoskeleton robot. Patients underwent training for the self-modulation of contralesional M1 excitability, supported by the closed-loop system, where somatosensory and visual stimuli were provided contingently based on the excitability of the targeted hemisphere. Our findings demonstrated that the intervention systematically induced targeted functional neural remodeling of the motor cortical network in post-stroke patients with severe motor disability and systematic improvement in upper-extremity motor abilities.

We adopted a single-center, single-arm, open-label, prospective trial design with eight patients to confirm the safety and preliminary clinical efficacy of the targeted neural remodeling approach designed to promote contralesional primary motor cortex (M1) excitability in severe chronic post-stroke hemiplegic patients, especially those with impaired shoulder mobility. From October 10, 2016 to May 10, 2017, at the Keio University Hospital, eight post-stroke patients (4 males/4 females) participated in the study with a mean age of 58.4 years (SD 11.1) and median time from stroke onset of 29 months (range, 14 to 97 months) (Supplementary Table 1). Recruited patients exhibited severe hemiplegia, as indexed by the average Fugl-Meyer Assessment (FMA, 16.75±4.4) at the pre-intervention evaluation.

The intervention was designed to promote functional remodeling of neural pathways originating from the contralesional M1 when patients attempted to elevate the paralyzed shoulder (Fig. 1a). To this end, we employed closed-loop brain-machine interaction training that was configured to activate the robotic device, functional electrical stimulation (FES), and virtual object movement contingent on the contralesional sensorimotor activities monitored by scalp EEG (Extended Data Fig. 1, Supplementary Movie 1). During training, patients were asked to elevate the shoulder on the affected side and self-modulate the bar height on the screen. The visual feedback represented contralesional M1 excitability evaluated by changes in the spectral power of the sensorimotor rhythm (SMR) in the scalp EEG^8^. Because SMR power attenuation reflects the excitability of neural populations in M1 and is referred to as event-related desynchronization (SMR-ERD)^9^, the robotic device and FES were activated when sustained SMR-ERD was observed for more than 1 second during the motor attempt period (Fig. 1b). The exoskeleton robot mimicked the movement performed by the occupational therapist and assisted shoulder flexion with FES.

**Fig. 1.**
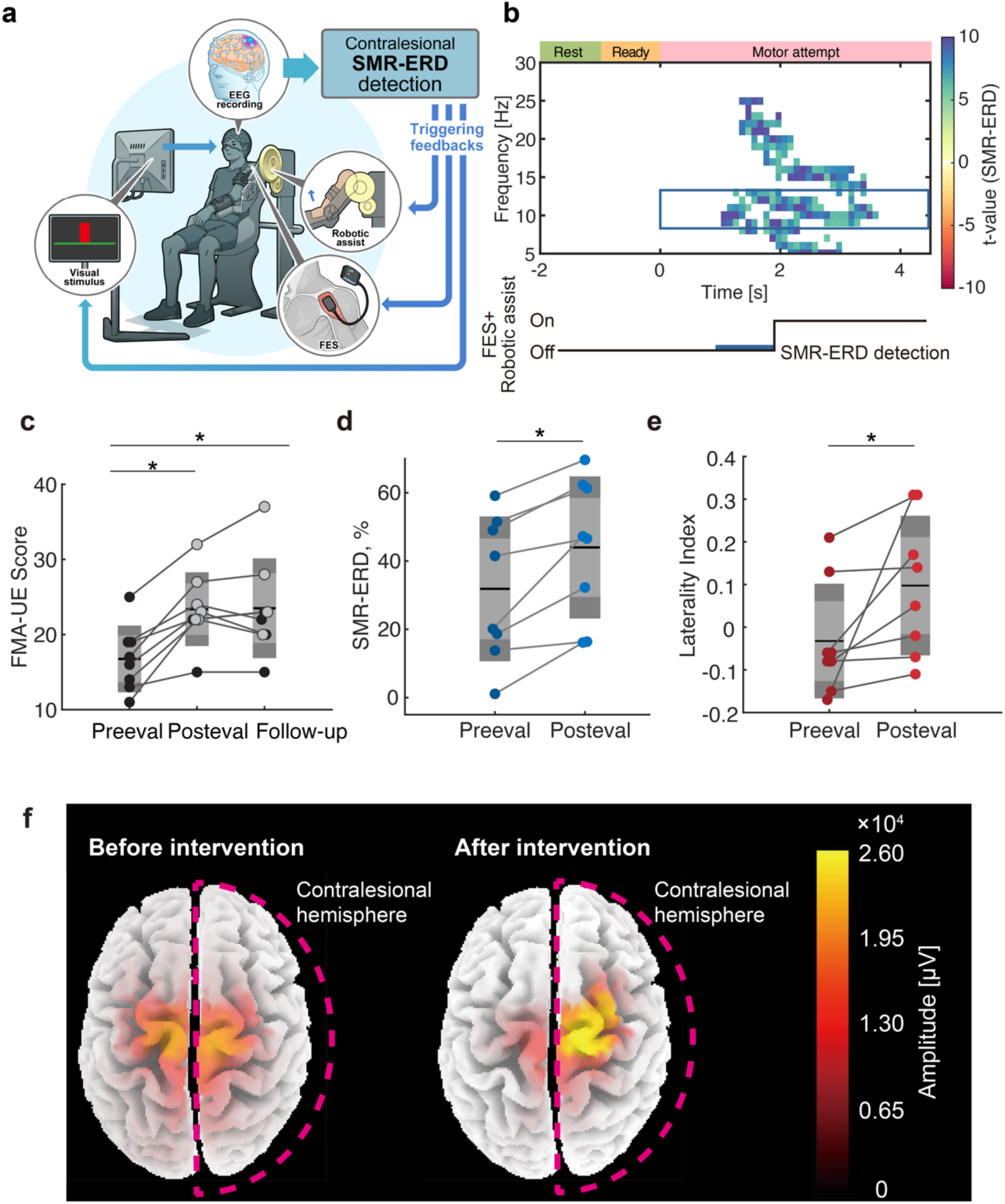
Brain-machine interaction training improves clinical scores and manipulates cortical sensorimotor representation. **a**. Schematic of the intervention. When patients attempt elevation of their affected arm, contralesional sensorimotor activities are online visualized as the bar height. When continuous was observed, robotic assist and electrical stimulation reconstructed arm elevation to mimic sensory feedback. EEG: electroencephalogram; SMR-ERD: event-related desynchronization of sensorimotor rhythm; FES functional electrical stimulation. **b**. Time course of the intervention and time-frequency representation of the contralesional scalp EEG data. Exoskeleton robotics assistance was initiated when SMR-ERD lasted for 1 s. **c**. Changes in FMA scores. Participants exceeding MCID were colored with light gray. FMA: Fugl-Meyer Assessment **d**. Changes in SMR-ERD magnitude. **e**. Changes in SMR-ERD laterality. **f**. Whole-brain representation of SMR-ERD, estimated by cortical source estimation algorithm (see methods) in the representative participant (Patient 4 in Table 1). The lateralization of SMR-ERD magnitude to the contralesional hemisphere is mainly observed around the sensorimotor cortex. One-way ANOVA with post-hoc Bonferroni-corrected *t*-test was conducted (*: *p* < 0.05).

Before and after a seven-day intervention as well as one-month follow-up period, we examined the upper limb ability. The primary outcome measure, FMA of upper extremity (FMA-UE), significantly improved after the intervention (Fig. 1c, One-way ANOVA, *p* < 0.0001, *F* = 20.854). All patients exhibited an increase in the FMA score after the intervention (6.6 ± 3.2), and 6 of 8 patients exceeded the minimal clinically important difference (MCID) value, which was set to 5 based on the initial FMA score^10^. The rest of the patients exhibited smaller yet systematic improvement. Changes in the scores were driven by the section for shoulder and wrist joint assessment (Extended Data Fig 2, Supplementary Movie 2). The FMA-UE score at the one-month follow-up period indicated five patients maintained the improved motor function exceeding MCID^10^. The rest of the patients exhibited sustained improvement relative to the pre-evaluation period. In addition, muscle tone indexed by Stroke Impairment Assessment Set^11^ (SIAS) was also improved in five patients, resulting in a significant increase between baseline and after the intervention as well as between baseline and one month after the intervention (Dunnett multiple comparison, *p* < 0.0001). Throughout the period, no adverse events were observed. Collectively, we found the efficacy and safety of the present intervention.

To test whether the observed clinical improvement is accompanied by the functional remodeling of the therapeutic target, we assessed the effects of this neurophysiological treatment on the contralesional M1. An analysis of the magnitude of contralesional SMR-ERD and its laterality revealed that SMR-ERD significantly increased after the intervention (Wilcoxon signed-rank test, *p* = 0.0120) and lateralized to the contralesional M1 (Wilcoxon signed-rank test, *p* = 0.0120) (Fig. 1d and e). These results suggest that the intervention induces contralesional SMR-ERD, as intended (Fig. 1f), ultimately allowing patients to learn to self-modulate contralesional M1 excitability.

Furthermore, we explored the whole-brain reorganization induced by the intervention. To this end, we tested the group-level changes in resting-state functional connectivity (rsFC) patterns using the imaginary part of coherency (iCOH) as a connectivity metric^12^. Significant increases in iCOH were found in the intra-hemispheric connection around the contralesional M1 (Fig. 2a), within the subject-specific frequency band. The intra-hemispheric network intensity, the sum of iCOH within the contralesional hemisphere, significantly increased while no significant changes were observed in other conditions (Fig. 2b). Collectively, three scalp EEG metrics consistently indicated that functional reorganization of cortical activity is observed in the contralesional M1, where the closed-loop training targeted.

**Fig. 2.**
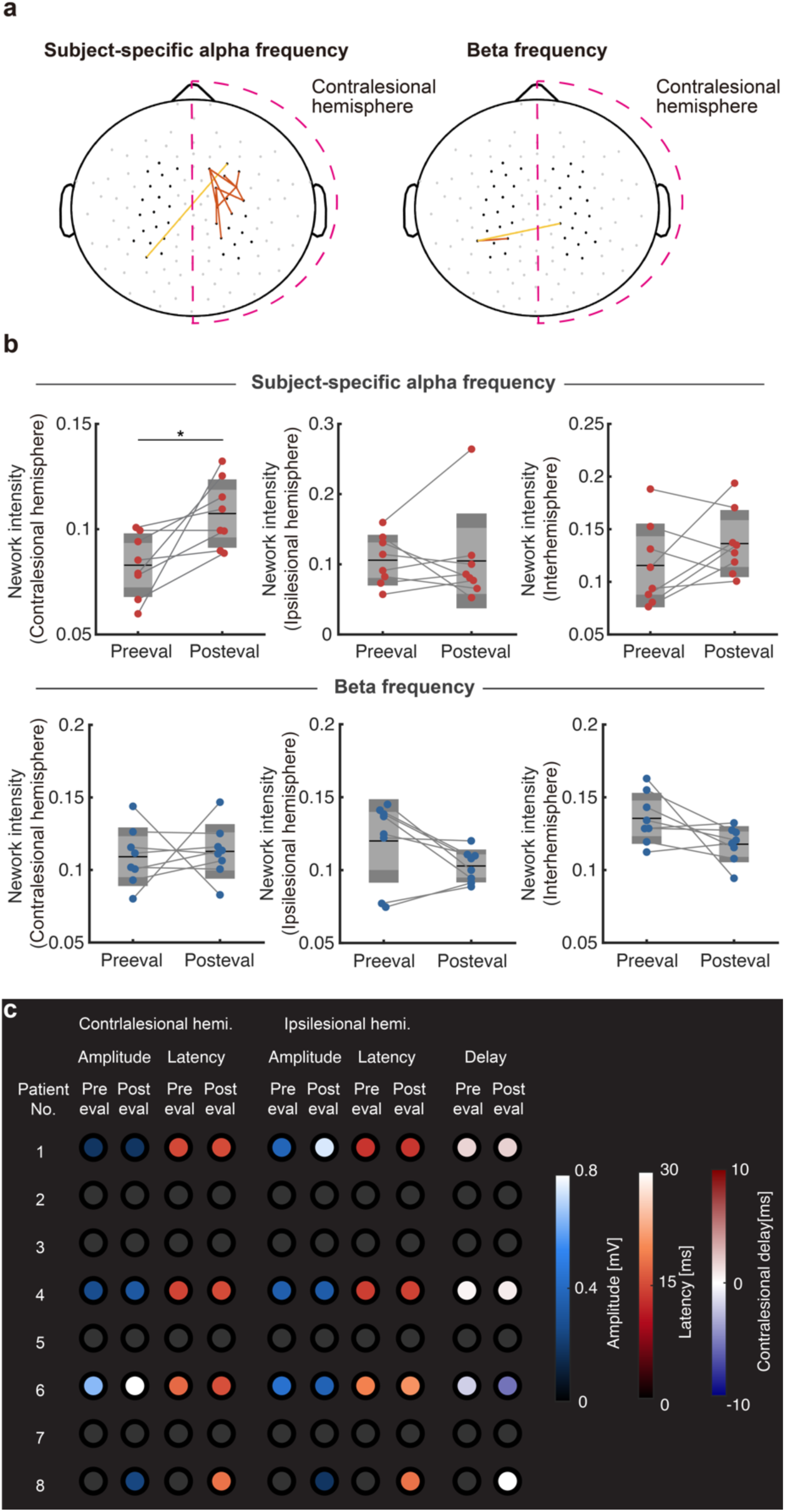
Induced sensorimotor plasticity revealed by resting-state connectivity patterns and MEP. MEP: motor-evoked potentials **a**. Statistically significant connectivity patterns in subject-specific frequency in the alpha- and beta bands estimated from the resting state EEG. Orange for intrahemispheric connections, and yellow indicates interhemispheric connections. **b**. Changes in the iCOH over contralesional and ipsilesional hemispheres in subject-specific alpha frequency (upper row) and beta band (lower row) for each patient. **c**. Changes in corticomotor responsiveness assessed by TMS. Each row of data represents one patient. The magnitudes of MEPs from the contralesional and ipsilesional hemispheres are represented with a color gradient from blue to white, while latencies are represented as a color gradient from red to white. Changes in latency are indicated as the delay in the contralesional hemisphere, with negative values signifying a faster response in the contralesional corticomotor pathway than in the ipsilesional pathway. TMS: transcranial magnetic stimulation

Finally, we tested whether the cortical reorganization observed in the scalp EEG metrics also influenced the corticomotor pathways directly connected with the affected muscle. We evaluated changes in the excitability of the corticospinal pathway from the contralesional M1 to the anterior deltoid muscle, an agonist muscle of shoulder elevation, using motor-evoked potentials (MEPs). Excitability was evaluated one day before and one day after the intervention to capture intervention-induced functional plastic changes in the pathways. MEPs from the paralyzed anterior deltoid muscle were evoked using transcranial magnetic stimulation (TMS) during 20% of maximum isometric voluntary contraction. Six patients without TMS contraindications were tested, and data from four patients who completed the evaluation were analyzed. Contralesional MEPs became apparent after the intervention in three of the six tested participants (Fig. 2c, Patients 4, 6, and 8). Patient 8, who exhibited no MEP before the intervention (Extended Data Fig. 3a), demonstrated reappearance, and within-participant statistical tests revealed that MEPs derived from the contralesional hemisphere of Patients 4 and 6, and the ipsilesional hemisphere of Patient 1, exhibited significant increases (two-sample t-test, Patient 4: *p* = 0.009, *d* = 1.07; Patient 6: *p* < 0.001, *d* = 3.30; and Patient 1*: p* < 0.001, d=3.29, Extended Data Fig. 3b). MEP latencies were maintained after the intervention in all tested patients.

Moreover, the potentiated corticomotor excitability from the contralesional side indicated that the affected muscle is voluntarily controlled via the motor pathway from the contralesional M1. Since these metrics reflect the effects of our intervention at both cortical and corticospinal levels, these analyses allow us to interpret the neurophysiological effects of the intervention. The results of electrophysiological assessments consistently indicate that our seven-day intervention rapidly reorganized the targeted brain region.

Here, we aimed to demonstrate that the noninvasive digital connection between the affected muscle and the intact contralesional M1 promotes functional remodeling of the targeted cortical areas and ipsilateral uncrossed pathways. We found that a seven-day consecutive use of our intervention manipulated the targeted neural pathways and led to functional motor recovery in post-stroke patients with chronic severe hemiplegia for whom it was determined that additional standard exercise would not overcome the ceiling effect. Moreover, the improvement in upper limb ability, as measured by FMA-UE scores, was greater than the MCID in six out of eight participants (FMA-UE MCID = 5)^10^. Given that impairments in motor function are treatment-resistant in patients with chronic severe hemiplegia, this single-arm, proof-of-concept study indicates the feasibility, safety, and efficacy of targeted neural remodeling of the intact contralesional corticospinal pathways using closed-loop interactions with this system.

The improvement of FMA-UE was mainly driven by the shoulder joint section (section A), where our treatment targeted, and partly by the wrist joint section at the follow-up period (section B). This remote effect of functional recovery may stem from use-dependent plastic changes induced by arm joint improvement and resource allocation of the arm area to the ipsilesional side by our intervention. As a result, patients might have exhibited the functional improvement of the wrist, due to unmasking cortical areas in the ipsilesional hemisphere that could contribute to distal part recovery.

We posit that it is a promising result that six out of eight participants at the chronic stage (1-8 years from stroke onset) exceeded FMA-UE gain more than MCID, in light of the previously reported recovery trajectory, specifically that patients achieve an FMA-UE of approximately 20 at 30 weeks from stroke onset^13–15^. According to the data from Winters et al^14^, all patients whose FMA scores were below 20 after 26 weeks had reached a plateau, and typically, no FMA changes were observed in this group. This observation suggests that patients in our cohort would not show any improvement if the treatment was not effective. Furthermore, data from pools with rehabilitation sessions once a week or more often, even during the chronic phase, indicated that our one-week period of daily intervention could induce recovery equivalent to that within the 6- to 12-month period after onset^13^, ultimately shortening the recovery period. Collectively, because the standard rehabilitation results in a plateau in upper-extremity ability at this period, it can be considered that the results observed in the current study were induced by our intervention. Therefore, by comparison with historical datasets, it can be inferred that our intervention significantly induced functional recovery of chronic, severe upper limb functional impairment compared to interventions affecting the standard locus. Further research, in which a direct comparison is performed under a controlled study design, is needed to obtain proof-of-concept data.

To examine whether our intervention successfully modulated the targeted neural structures as designed, we sought the neurophysiological signatures of the functional remodeling of the contralesional M1. The contralesional SMR-ERD magnitude was the target of self-regulation during the intervention and successfully lateralized across participants. It suggests that the intervention successfully induced the hemispheric activation of the contralesional hemisphere. Related, rsFC change over the contralesional hemisphere, was mainly found in the frequency used for machine actuation during the intervention, especially for the contralesional M1 and premotor areas and their surrounding channels. Since these channels are positioned near M1 and the premotor cortex of the right hemisphere, the observed FC changes induced by our intervention might reflect the reinforced connectivity network among M1 and the premotor cortex in the contralesional hemisphere. In line with the present finding, motor recovery has been reported to be accompanied by the reorganization of the sensorimotor network^16^ and a significant increase in intrahemispheric connections in the contralesional hemisphere for patients who achieved good recovery after stroke^17^. Given that a positive contribution of the contralesional cortical network for learning novel motor skills was found in post-stroke hemiplegic patients, these network changes may underlie the successful control of excitability in the target region during the intervention and motor recovery.

In addition to the EEG signatures, we examined the small number of patients in TMS experiments to further evaluate the corticospinal excitability. In the group, three of four tested participants showed increased ipsilateral MEPs, and 1 showed the reappearance of ipsilateral MEPs. Because studies have reported a positive correlation between MEPs and SMR-ERD over the contralateral and ipsilateral hemispheres in response to unilateral motor imagery^18,19^, the self-modulation training of contralesional SMR-ERD may have induced plastic changes in the contralateral M1. However, due to the limited number of patients in this pilot TMS assessment, further studies on such neurophysiological reorganization are required.

There were several limitations of this study. First, the sample size to test the primary safety and efficacy of the intervention was small by design. Thus, the generalizability of the findings needs to be clarified. Furthermore, while the patients were blinded to targeted channel location and the training parameters, the clinicians were not blinded; this may have influenced the clinicians’ interpretation of the therapeutic response. It is also possible that the combination of repetitive electrical stimulation with the exoskeleton robot elicited motor recovery, independent of the brain-state dependent configuration. We posit that this recovery is unlikely because recruited participants had undergone standard clinical treatment and showed no significant response before enrollment in this study. Since we confirmed the feasibility, safety, and preliminary efficacy of treatment under these limitations, further controlled studies would reveal the clinical significance of the brain-machine interaction therapy.

In the present study, we found that the closed-loop, brain state-dependent machine actuation promoted the functional remodeling of the contralesional M1 in patients with chronic severe post-stroke hemiplegia. We posit that this intervention would be beneficial even for patients whose EMG data do not exhibit a response and whose active range of motion of the upper extremities is limited. After our intervention, these patients could subsequently experience behavioral training, such as EMG feedback^20^. Because such a sequential combination of neural intervention and motor exercise induces a cumulative therapeutic effect^20^, the digital therapeutic approach can be combined with standard clinical treatment to expand the range of treatable conditions.

---

**Extended Data Fig. 1.**
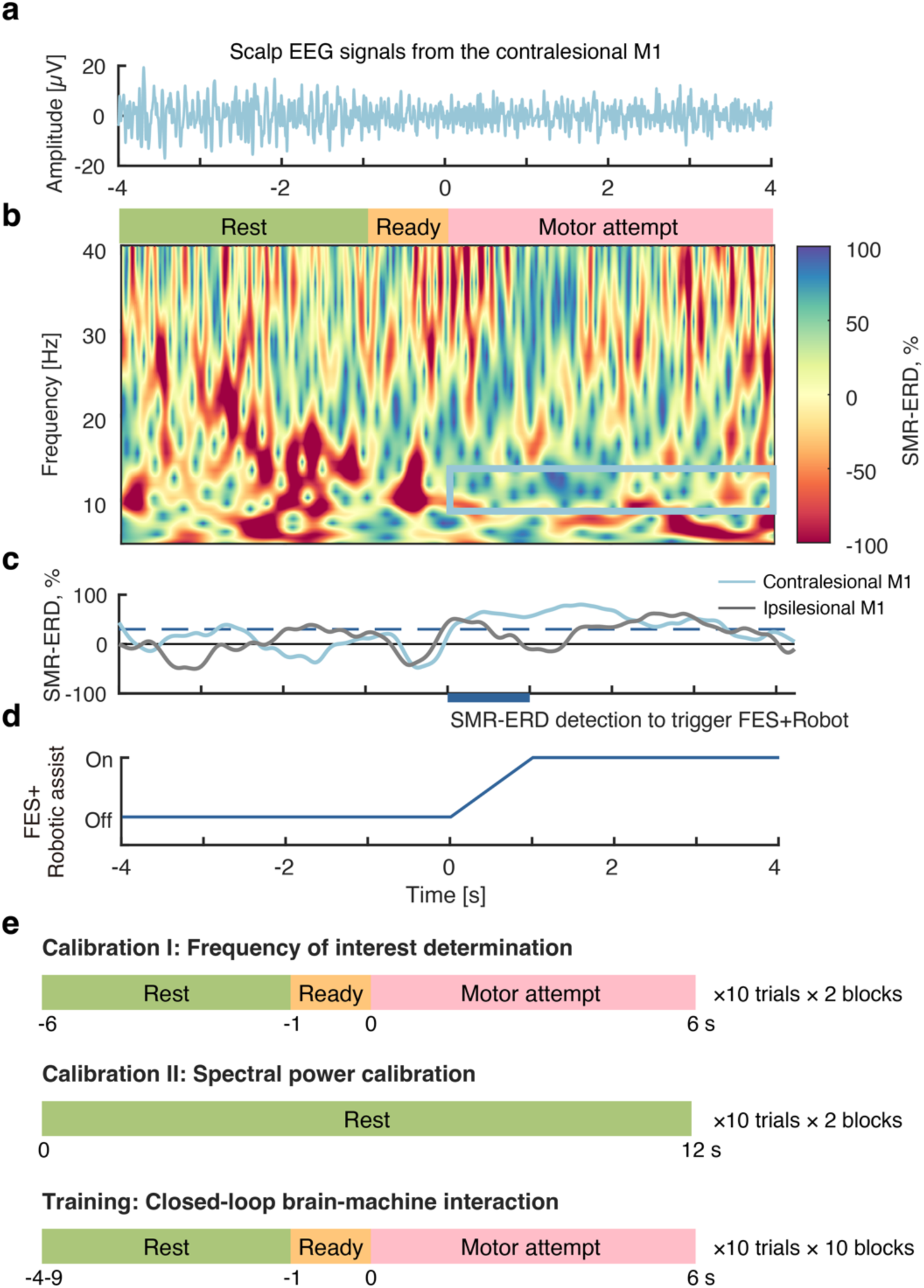
Action sequence and trial structures of the intervention procedure. **a**. raw scalp EEG signals derived from the contralesional motor cortex (M1). **b**. Time-frequency representation of EEG signals from a single trial. The blue square indicates targeted frequency used for the SMR-ERD calculation. **c.** time course of SMR-ERD magnitude from the contra- and ipsi-lesional hemisphere. **d**. time course of electrical stimulation and robotic assist. Machines are actuated when sustained SMR-ERD magnitude was observed for one second. **e.** Trial structures in calibration and training sessions.

**Extended Data Fig. 2.**
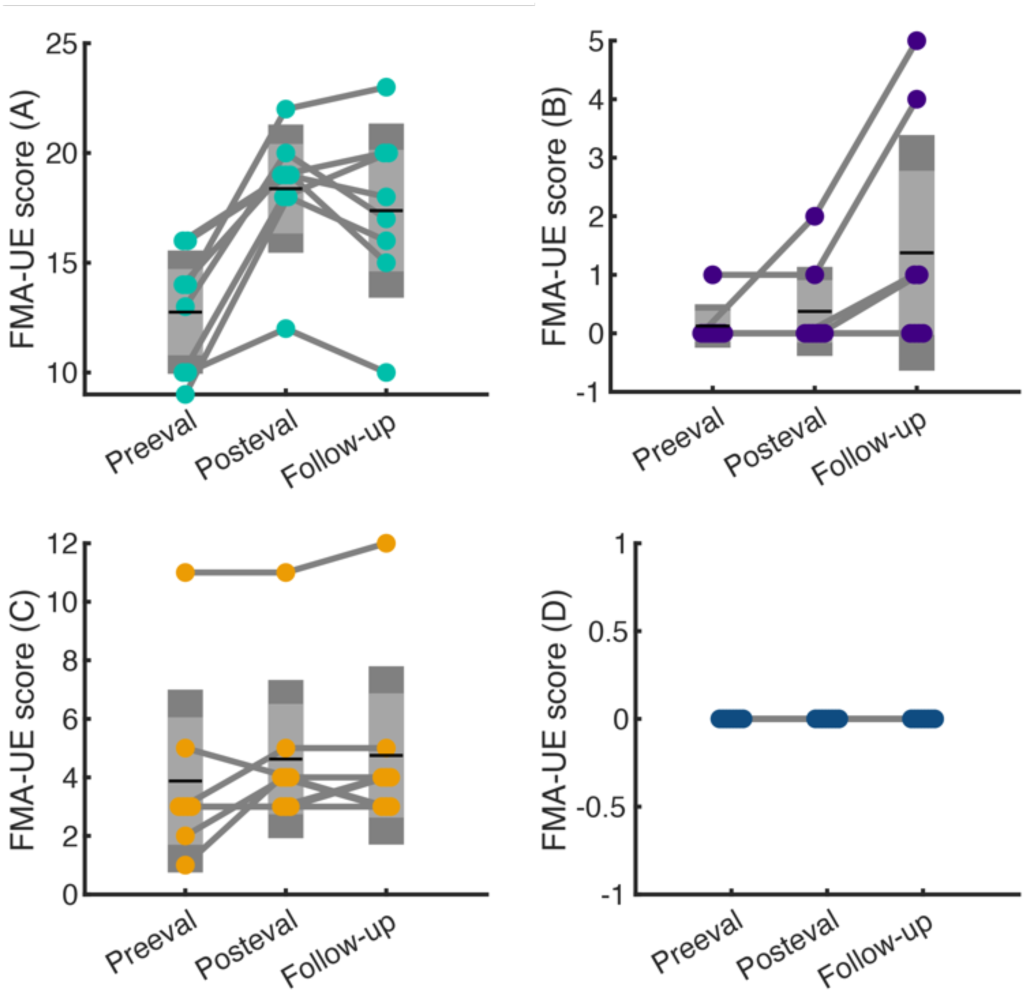
Subscores of FMA Scores. The subscores of A, B, C and D represents shoulder, wrist, hand joint function and their coordination, respectively.

**Extended Data Fig. 3.**
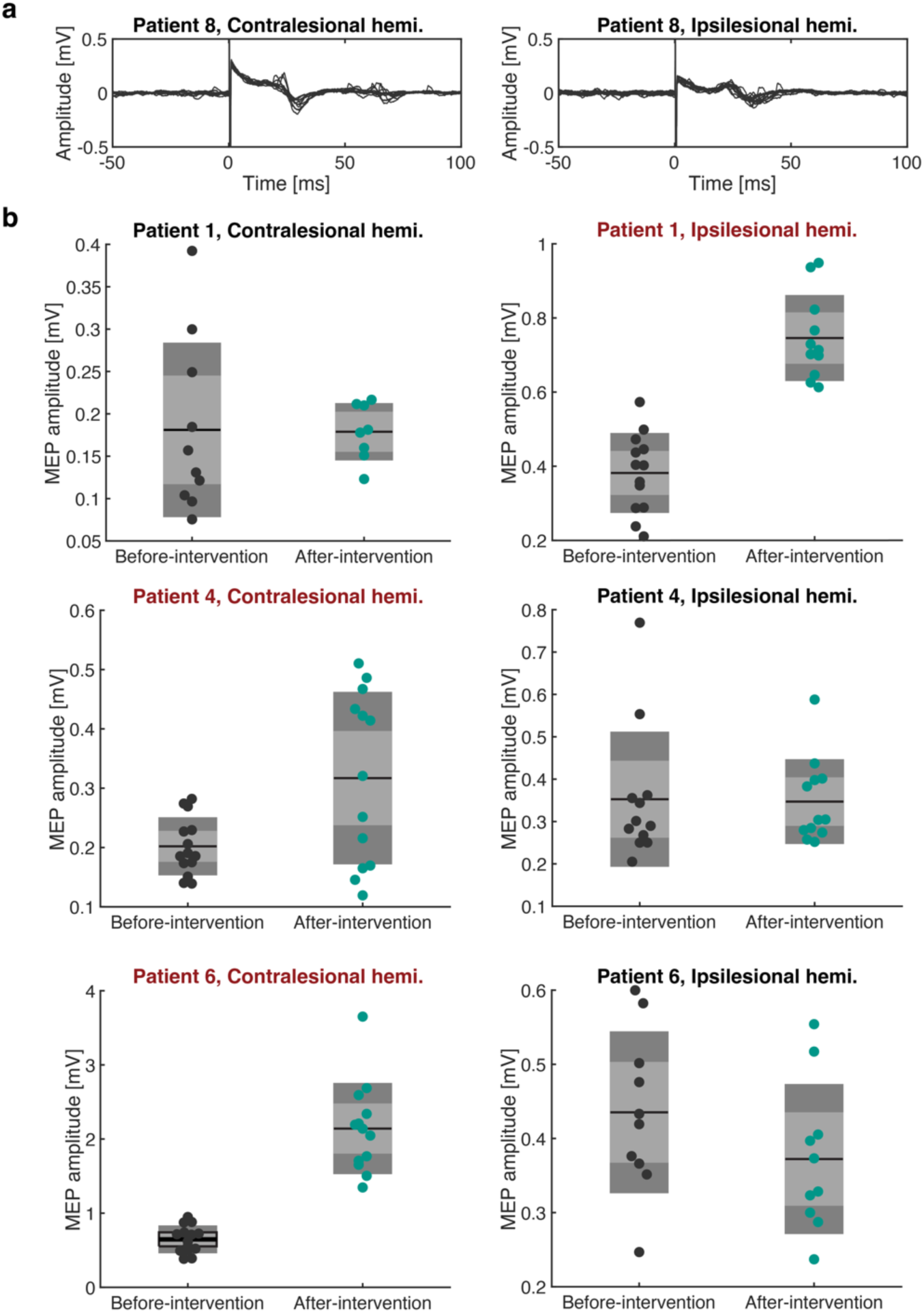
The TMS assessment reveals that functional remodeling of the corticospinal pathways was induced by the intervention. **a.** Raw signals of MEP from a representative participant at after the intervention period. Left and right panel indicate MEP from anterior deltoid muscle derived by stimulations on the contralesional and ipsilesional hemisphere, respectively. **b.** MEP magnitudes each participant. Data in each panel were subjected to two-sample t-tests with Bonferroni correction for each hemisphere. Titles whose data exhibited significant difference between before and after intervention were colored with red.

## Conflicts of Interest

J.U. is a founder and representative director of the university startup company, LIFESCAPES Inc., which is involved in the research, development, and sales of rehabilitation devices, including brain-machine interfaces. He receives a salary from LIFESCAPES Inc. and holds shares in LIFESCAPES Inc. S.I receives a salary from LIFESCAPES Inc. This company does not have any relationships with the device or setup used in the current study. The remaining authors declare no competing interests.

## Acknowledgments

This study was partially supported by the Strategic Research Program for Brain Sciences from the Ministry of Education, Culture, Sports, Science, and Technology of Japan Grant Number 15H05880 and JSPS KAKENHI Grant Number 20H05923, Japan.

## Funding Source

The study’s sponsors had no role in the study design, data collection, data analysis, data interpretation, or writing of the report. All authors had full access to all the data in the study, and the corresponding author had the final responsibility of the decision to submit for publication.

## Methods

### Ethics

The intervention was conducted in accordance with the Template for Intervention Description and Replication (TIDieR) guidelines^21^ and the CONSORT extension for pilot and feasibility studies^22^. The institutional ethics review board at Keio University School of Medicine approved the study (approval 20140442 and 20241023), registered with the University Hospital Medical Information Network (UMIN) (UMIN000017525). The experiment was performed after written informed consent was obtained from the participants and conducted in accordance with the Declaration of Helsinki (Trial Registry: UMIN Clinical Trials Registry; No. UMIN000017525.)

### Study Design and Participants

We adopted a single-center, single-arm, open-label, prospective trial with a limited number of patients to confirm the safety and preliminary clinical efficacy of the closed-loop brain-machine intervention designed to promote contralesional M1 excitability in severe chronic poststroke hemiplegic patients^23^.

The total sample size was set as 8 based on the following rationale: the minimal detectable change in the Fugl-Meyer Assessment (FMA) in chronic poststroke hemiplegic patients is reported to be 3.2 points^24^, and we aimed to test whether the intervention effectively induces detectable FMA gains. Therefore, given that the estimated Cohen’s *d* (effect size) for robot therapy in 66 patients was 1.05 (FMA change: 3.80 ± 3.6 points), the required sample size for the paired-sample *t* test to detect this effect size in the pre- to postevaluation comparison was estimated to be 8.

Participants were recruited from stroke outpatients at Keio University Hospital with the goal of selecting chronic stroke patients (to avoid spontaneous recovery as a confounder) with severe motor deficits in the upper limbs, especially the shoulder. The inclusion criteria were as follows: (i) first unilateral, cortical, subcortical, or mixed stroke; (ii) time from stroke onset longer than 180 days; (iii) age between 20 and 80 years; (iv) knee-mouth test score on the SIAS less than two, which represents a patient can only lift the hand to the level of the nipple^11^; (v) passive range of motion more significant than 90 degrees for affected shoulder flexion; and (vi) ability to walk independently in daily life with or without assistance. Exclusion criteria were the presence of (i) severe cognitive deficits, such as unilateral spatial neglect or aphasia, precluding training; (ii) severe proprioceptive deficits, muscle tone reduction, or pain in the paretic upper extremity, as measured by the Stroke Impairment Assessment Set^11^ (SIAS); (iii) pacemaker or other implanted stimulator; and (iv) other severe medical conditions. Finally, eight poststroke hemiplegic patients (4 males/4 females) were recruited (see supplementary information for detailed characteristics).

### Safety Assessment

Occupational and physical therapists (M.O and K.O) designed the safety assessment, and rehabilitation doctors (F.L and M.K) checked the intervention risks according to the ISO14971 stroke rehabilitation guidelines;^25^ if necessary, risk countermeasures were applied before the intervention. The safety outcome measure was the occurrence of one of five adverse events: shoulder pain, abrasion, heat burn by electrical stimulation, ischemia, and infection. These items represent the safety concerns associated with passive movement training of the shoulder joint. The safety assessment was peer-reviewed by two rehabilitation doctors, one occupational therapist, and one physical therapist and performed by one occupational therapist and one physical therapist. Participants were queried about adverse events at each study contact.

### EEG Data Acquisition

Scalp EEG signals were recorded with a 128-channel Geodesic EEG system (EGI, Oregon, United States) covering the whole head. Electrode impedance was kept below 40 kΩ throughout the experiment. The EEG signals were amplified and filtered between 0.01 and 70 Hz and sampled at 1000 Hz; power noise was also removed. Participants were instructed to keep their arms and hands relaxed during recordings.

### Online EEG Data Processing

The excitability of the contralesional motor cortex was evaluated based on changes in the spectral power of the sensorimotor rhythm by scalp EEG. The power attenuation of SMR is thought to reflect the excitability of populational activities in M1^18^ and is referred to as event-related desynchronization of SMR (SMR-ERD). The SMR-ERD magnitude was determined relative to EEG power in a specific frequency band, such as the alpha (8-12 Hz) or beta (12-20 Hz) band. In these frequency bands, attenuation of spectral power was observed during motor-related tasks^26^. In this study, the SMR-ERD magnitude derived from the contralesional M1 was calculated during training using the following formula:

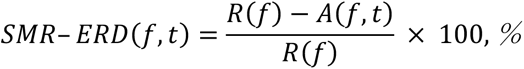

where *A* is the power spectrum density of the recorded EEG at frequency *f* and time *t* regarding the onset of motor imagery, and *R* is the power spectrum density of a 2-second period from 1 second after the beginning of rest to 1 second before the end of the resting period. Moreover, we selected the individual alpha and beta frequencies for both online and offline analyses to capture individually variable responsive frequency bands. The frequency band range was set to 2 Hz in 8-20 Hz (alpha + beta), as the most significant SMR-ERD magnitude was obtained in the 2 Hz frequency band during calibration sessions^27^.

EEG data of each trial was collected to calculate SMR-ERD magnitude in the following procedure based on the previous closed-loop intervention for finger motor recovery^3,17^: (1) common averaged filter, bandpass (0.01−70 Hz), and notch (50 Hz) filtering with a fourth-order Butterworth filter, (2) segmentation for 1 s of EEG data with 90% overlap, (3) Fast Fourier transform (FFT) with Hann window function, (4) power spectrum density (PSD) calculation in participant’s specific frequency bands, (5) SMR-ERD calculation.

### Intervention Procedures

The intervention consisted of a two-block calibration period and a four-block training session. Each block lasted approximately two minutes. In total, participants underwent 30 minutes of intervention per day (excluding the intermission and EEG cap setup time). The interventions were carried out for seven consecutive days without conventional exercise. Clinical and neurophysiological measurements were performed one day before and after the intervention. A custom-made exoskeleton robot (Advanced Telecommunications Research Institute International) was attached to the paralyzed arm, and the other arm was kept relaxed throughout the intervention. The exoskeleton robot mimicked the movement performed by the occupational therapist and assisted shoulder flexion with functional electrical stimulation (FES) with the following parameters: frequency: 100 Hz, pulse width: 1 ms, intensity: motor threshold. Proprioceptive feedback was provided in the form of movement assistance during the intervention using the shoulder exoskeleton driven by a pneumatic cylinder^3,28,29^. A monitor was placed 60–90 cm in front of the participants to present visual instruction and feedback about the current brain state.

The calibration session consisted of two different parts: the first part served to identify the EEG frequency band of interest for both online and offline analyses, and, in the second part, the baseline distribution of SMR power^17^ was calculated. Each part consisted of 2 runs of 10 trials, for a total of 20 trials. In the first part, participants were asked to attempt shoulder flexion at regular intervals. Each trial started with a 5-second resting epoch, followed by a 1-second preparation epoch, and ending with a 6-second motor attempt epoch. In the second part, participants were asked to relax for the 12 second duration of each trial.

In the training sessions, participants were asked to attempt shoulder flexion (Extended Data Fig. 1). Training consisted of 10 blocks of 10 trials, for a total of 100 trials. Each trial started with a resting period that lasted 4–9 seconds, followed by a 1-second preparation period and finally a 6-second MI period. The resting period finished when the SMR power was within 1 SD of the mean estimated in the calibration session. If the power did not meet the criteria, the trial ended without going to the next period. During the task period, visual SMR-ERD feedback was presented. When the participants maintained SMR-ERD over 30% for 1 second, robotic assistance with shoulder flexion and FES was triggered (Fig. 1a).

### TMS

To evaluate the effects of the intervention, changes in corticomotor excitability were tested using TMS. Evaluation was performed using a Magstim 200 magnetic stimulator (Magstim, Whitland, UK) with an angulated (95°) double cone coil one day before and after the intervention. The coil was optimally positioned to obtain MEPs in the anterior deltoid muscle with the lowest stimulus intensity, and the position was monitored with a Brainsight TMS navigation system (Rogue Research, Cardiff, UK). MEPs of the paralyzed anterior deltoid muscle were measured during shoulder flexion at 20% of maximum isometric voluntary contraction in 6 of 8 participants who did not have contraindications for TMS. Single-pulse TMS was applied over the contralesional (i.e., ipsilateral) and ipsilesional (i.e., contralateral) M1 at 110% active motor threshold. EMG activity from the paralyzed anterior-deltoid muscle was recorded with a pair of Ag/AgCl surface electrodes (10 mm diameter) placed with centers 20 mm apart over the muscle bellies. The amplified and bandpass-filtered (5-1 kHz) raw EMG signal was initially digitized at a 20 kHz sampling rate, downsampled at 5 kHz, and stored for later analysis of MEP amplitudes and latencies.

### Outcome Measures

The FMA of upper extremity (FMA-UE) score was the primary outcome measure since it represents limb impairment in terms of synergistic motor control^30^. The minimal clinically important difference (MCID) value was set to 5 based on the initial FMA score, adopting a conservative threshold. The SIAS was the secondary outcome measure for clinical testing; this standardized measure of stroke impairment consists of 22 subcategories and has excellent interrater reliability. Motor functions of the paretic upper extremity were evaluated with the knee-mouth test and the finger test and rated from 0 to 5, with 0 indicating complete paralysis and 5 indicating no paresis. The MCID was set at 1 point, indicating that a visual change occurred. Both FMA and SIAS were measured approximately one month after intervention in a follow-up test.

The secondary outcome measures of the neurophysiological assessment consisted of the following established metrics: (1) MEP size derived from the contralesional hemisphere, (2) SMR-ERD magnitude used as a biomarker of M1 activity^3^, (3) laterality of SMR-ERD^31^, and (4) resting-state functional connectivity (rsFC) of whole-head scalp EEG signals^17,32^. We performed TMS to quantify changes in excitability in neural pathways from the M1 to the ipsilateral muscles.

### Neurophysiological Analysis -scalp EEG-

To evaluate the target specificity of the intervention designed to train contralesional hemispheric modulation, the laterality index (*LI*) of the SMR-ERD magnitude was calculated using the following formula:

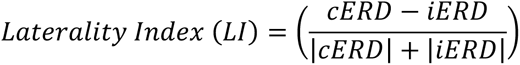

where cERD and iERD are the values of the contra- and ipsilesional ERD, respectively.

Given that instantaneous neural oscillatory phase patterns reflect the efficacy of neuronal communication, rsFC is thought to reflect how spatially and spectrally targeted upconditioning training changes cortical network properties. To comute rsFC from scalp EEG data, the imaginary part of coherence (iCOH) has been described as a promising analysis to reveal underlying connectivity, and iCOH-based FC measures have been provided by recent research^12,33^. Accordingly, lagged coherence, which has been developed to give an improved connectivity measure compared to the iCOH, is one of the iCOH-based connectivity measures that has been used in sensor-level EEG connectivity analysis^32^. Since EEG signals measured by our high-density sensors are sensitive to volume conduction effects, we adopted iCOH as sensor-level rsFC measures, which are robust to these effects^34^. The iCOH was estimated from resting-state EEG data to determine the change in intrahemispheric rsFC in both ipsilesional and contralesional hemispheres and that of interhemispheric rsFC after the intervention. iCOH between EEG channels *i* and *j* was calculated with the following procedure. First, their cross-spectrum was calculated as

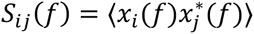

where *x_i_*(*f*) and *x_i_*(*f*) are the corresponding Fourier transforms of signals obtained from EEG channels *i* and *j*, respectively. Then, the complex-coherence is computed as *S_ii_ S_jj_*

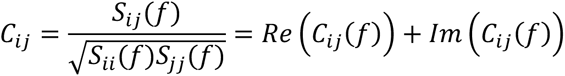

where *S_ii_*(*f*) and *S_jj_*(*f*) are the auto-spectrum of channels *i* and *j*. Lagged coherence is then calculated as the normalized imaginary part of coherence using the real part of coherence:

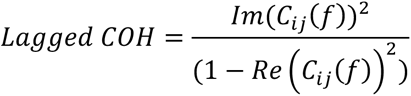

To test whether relevant network connectivity significantly increased after the intervention, we created amplitude-matched surrogate data and calculated the 95^th^ percentile as the threshold for detecting significant FC for each pair of channels. Intra- and interhemispheric network connectivity was estimated as the average lagged COH between the channels of a given hemisphere. We selected 34 channels, excluding the peripheral channels and those that detected eye movement artifacts in our connectivity analysis. The frequency of interest was set to the subject-specific frequency band, and the beta band was considered as a control condition.

### Statistical Analysis

We used repeated measures one-way ANOVA with a post hoc Dunnett’s multiple comparisons test to evaluate FMA-UE scores at baseline, post intervention (On the next day of the last intervention), and one month after the intervention.

Changes in SMR-ERD magnitude in the contralesional hemisphere and their LI values were analyzed with the Wilcoxon signed-rank test. To determine the significant FC estimated at the sensor level, resting-state EEG data were evaluated using surrogate data^35^. The distribution of surrogate data was used to estimate the threshold at the 95^th^ percentile for detecting significant FC in each patient. Then, a one-tailed paired-sample *t* test with FDR correction was performed to determine if significant increases in iCOH were observed at the group level. For MEP data, two-sample *t*-test was applied for data from each participant. For all statistical tests, significance threshold was set as 0.05.

## Data Availability

Supplementary movies are available upon request from the corresponding author.

## Notes

### Clinical Trial

UMIN000017525

### Funding Statement

This study was funded by the Strategic Research Program for Brain Sciences from the Ministry of Education, Culture, Sports, Science, and Technology of Japan Grant Number 15H05880 and JSPS KAKENHI Grant Number 20H05923, Japan.

### Author Declarations

The institutional ethics review board at Keio University School of Medicine approved the study (approval 20140442 and 20241023), registered with the University Hospital Medical Information Network (UMIN) (UMIN000017525).

