## Supplementary Information for "Rapid functional remodeling of the targeted contralesional hemisphere induced by one week of noninvasive closed-loop neurofeedback guides motor recovery in post-stroke patients with chronic motor impairment: a phase I trial"

**Table 1. Characteristics of the patients at baseline**

| Patient | Stroke Type | Damaged lesion | Age, y | Time since stroke, mo | FMA | SIAS KM | Proprioceptive deficit |
|---|---|---|---|---|---|---|---|
| 1 | Infarction | L CR | 66-70 | 19 | 25 | 2 | None |
| 2 | Haemorrhage | L TH | 51-55 | 26 | 17 | 2 | None |
| 3 | Haemorrhage | L PU | 46-50 | 31 | 19 | 2 | Mild |
| 4 | Infarction | R CR | 66-70 | 29 | 19 | 2 | None |
| 5 | Infarction | L M1 | 46-50 | 14 | 11 | 2 | None |
| 6 | Haemorrhage | L TH | 71-75 | 97 | 14 | 2 | Moderate |
| 7 | Infarction | R CR | 46-50 | 29 | 16 | 2 | None |
| 8 | Infarction | R CR | 61-65 | 34 | 13 | 2 | Mild |

R: right; L: left; TH: thalamus; CR: corona radiata; PU: putamen; M1: primary motor cortex; FMA: Fugl-Meyer Assessment; SIAS KM: Stroke Impairment Assessment Set Knee-Mouth Test

**Table 2: Averaged MEP amplitudes and latencies at pre- and post-intervention**

| Patient | Contrlalesional MEP [mv] | | Ipsilesional MEP [mv] | | Contrlalesional latency [ms] | | Ipsilesional latency [ms] | | contralesional delay [ms] | |
|---|---|---|---|---|---|---|---|---|---|---|
| | pre | post | pre | post | pre | post | pre | post | pre | post |
| 1 | 0.18 | 0.18 | 0.38 | 0.75 | 15.0 | 15.3 | 13.3 | 13.5 | 1.7 | 1.8 |
| 2 | - | - | - | - | - | - | - | - | - | - |
| 3 | - | - | - | - | - | - | - | - | - | - |
| 4 | 0.27 | 0.32 | 0.35 | 0.34 | 14.6 | 15.2 | 14.1 | 14.4 | 0.5 | 0.8 |
| 5 | - | - | - | - | - | - | - | - | - | - |
| 6 | 0.65 | 2.14 | 0.44 | 0.37 | 17.6 | 15.6 | 19.5 | 21.1 | -1.9 | -5.5 |
| 7 | - | - | - | - | - | - | - | - | - | - |
| 8 | N/A | 0.24 | N/A | 0.18 | N/A | 18.3 | N/A | 18.4 | N/A | -0.1 |

MEP: Motor evoked potential

| Table 3: FMA-UE scores | | | | | | | | | | | | | | | |
|---|---|---|---|---|---|---|---|---|---|---|---|---|---|---|---|
| Patient | A | | | B | | | C | | | D | | | Total | | |
| | pre | post | follow | pre | post | follow | pre | post | follow | pre | post | follow | pre | post | follow |
| 1 | 14 | 19 | 20 | 0 | 2 | 5 | 11 | 11 | 12 | 0 | 0 | 0 | 25 | 32 | 37 |
| 2 | 16 | 19 | 18 | 0 | 0 | 1 | 3 | 3 | 4 | 0 | 0 | 0 | 19 | 22 | 23 |
| 3 | 10 | 18 | 20 | 0 | 0 | 0 | 1 | 4 | 3 | 0 | 0 | 0 | 11 | 22 | 23 |
| 4 | 14 | 22 | 23 | 0 | 0 | 0 | 3 | 5 | 5 | 0 | 0 | 0 | 17 | 27 | 28 |
| 5 | 16 | 19 | 15 | 1 | 1 | 4 | 2 | 4 | 3 | 0 | 0 | 0 | 19 | 24 | 22 |
| 6 | 13 | 20 | 17 | 0 | 0 | 0 | 3 | 3 | 3 | 0 | 0 | 0 | 16 | 23 | 20 |
| 7 | 10 | 12 | 10 | 0 | 0 | 1 | 3 | 3 | 4 | 0 | 0 | 0 | 13 | 15 | 15 |
| 8 | 9 | 18 | 16 | 0 | 0 | 0 | 5 | 4 | 4 | 0 | 0 | 0 | 14 | 22 | 20 |

FMA: Fugl-Meyer Assessment
